# “Novel Clinical Manifestations of Human Monkeypox Virus Infection and Current Therapeutic and Preventive Strategies: A Systematic Review “

**DOI:** 10.1101/2023.01.06.23284258

**Authors:** Santenna Chenchula, Mohan Krishna Ghanta, Madhavrao Chavan, Krishna Chaitanya Amerneni, R Padmavathi, Rupesh Gupta

**Affiliations:** Department of Pharmacology, All India Institute of Medical Sciences, Bhopal, India; Department of Pharmacology, MVJ Medical College and Research Hospital, Bangalore, Karnataka; Department of Pharmacology, All India Institute of Medical Sciences, Mangalagiri, India; Faculty, Western Michigan University, Kalamazoo, Michigan; SVSMCH, Mahbubnagar, Telangana, India; Department of Internal Medicine, Government Medical College, Shahdol, Madhya Pradesh, India

**Keywords:** Monkeypox, Novel Antivirals, Monkeypox outbreak 2022, Tecovirimat

## Abstract

The escalating global monkeypox cases since early May 2022, acquired by the monkeypox virus (MPXV), a double-stranded DNA virus has been declared a public health emergency of international concern by the World Health Organization (WHO). Globally as of December 2022, the MPXV was transmitted to more than 100 countries with around 82,550 cases, among which 81577 cases from the 103 countries that are non-endemic to the MPXV with more than 50 deaths. The ripple effect of the Monkeypox outbreak in nonendemic countries globally could potentially bring significant challenges to worldwide health systems if the spread of the virus is not effectively controlled. In this urgent situation, only three antiviral drugs are in use against monkeypox infections and are not specific against monkeypox, hence the scientific communities across the world are in search to explore vaccines or therapeutic antiviral drugs selectively against the monkeypox virus. Here, in the present review, we discuss the clinical characteristics and treatment outcomes of the ongoing MPX outbreak of 2022 in nonendemic countries globally, from the published and grey literature in PubMed, Scopus and Google scholar. A total of 17 studies with 17,811 of MPX cases were found and included in the final qualitative analysis of the current systematic review.

## Background

In the present smallpox post-eradication era, monkeypox (MPX) has emerged as the most important and high-threat orthopox virus infection spread among humans [1]. Monkeypox *virus (MPXV)*, is the causative agent of the MPX infection, a double-stranded DNA virus with an envelope belonging to the orthopoxvirus genus of the family Poxviridae. Recently, the WHO renamed monkeypox as “mpox”. Since early May 2022, a rapid surge in the cases of monkeypox from more than 100 non-endemic countries and some endemic counties across the world is a worrisome and highly considerable public health issue [1]. Globally as of December 2022, the MPX virus transmitted to more than 100 countries with around 82550 cases, among which 81577 cases were from the 103 countries that are non-endemic to the MPX virus with more than 50 deaths from non-endemic countries have been reported current MPX outbreak [1-2]. The two common MPV transmissions are animal-to-human and human-to-human [3]. The WHO reported that in the past 30 years, monkeypox was endemic in several African countries with most suspected cases of MPX occurring in the Democratic Republic of the Congo till May 2022[1]. There are two distinct clades of monkeypox virus, the Congo Basin clade (endemic in Central Africa) and the West African clade [1-2]. On July 23, the WHO Director-General declared the escalating global monkeypox outbreak a Public Health Emergency of International Concern (PHEIC), highlighting the need for an effective and equitable response that includes accelerated access to diagnostic testing, vaccination and treatment. Because of the several animal reservoirs, complete eradication of MPXV is not possible [1].

To curtail the rapid spread and severity of MPXV epidemics in nonendemic regions of the world, it is necessary to have an in-depth understanding of the virology, transmission, clinical symptoms and management strategies of the current monkeypox outbreak 2022 reported cases of MPX infection. There is a dearth of information on the novelty of the current MPX outbreak clinical virology, transmission, signs and symptoms as well as treatment outcomes of MPX infection in non-endemic regions. Therefore, in the present review, we have analysed to identify the novel clinical characteristics of the current MPX outbreak reports across the world from the published and grey literature.

## Methodology

We conducted a systematic literature search from the 1^st^ May 2022 to December 2022 in the PubMed, Cochrane Library, Google Scholar databases and grey literature using the Mesh terms or keywords “Human Monkeypox” OR “Monkeypox” OR “Monkeypox Virus” OR “MPX” OR “MPXV”, to study the current MPX outbreak clinical characteristics and treatment outcomes in the non-endemic regions across the world. A comprehensive review of the published and grey literature on the current outbreak, to inform the novel clinical characteristics of the current Monkeypox outbreak and treatment outcomes in comparison to previously reported MPX infection clinical characteristics in the nonendemic countries across the world.

## Results

In the present review, we discuss the clinical characteristics and treatment outcomes of the ongoing MPX outbreak of 2022 in nonendemic countries globally, from the published and grey literature in PubMed, Scopus and Google scholar. A total of 17 studies with 17,811 of human MPX cases were found and included in the qualitative analysis [**Figure** 1].

**Figure 1:**
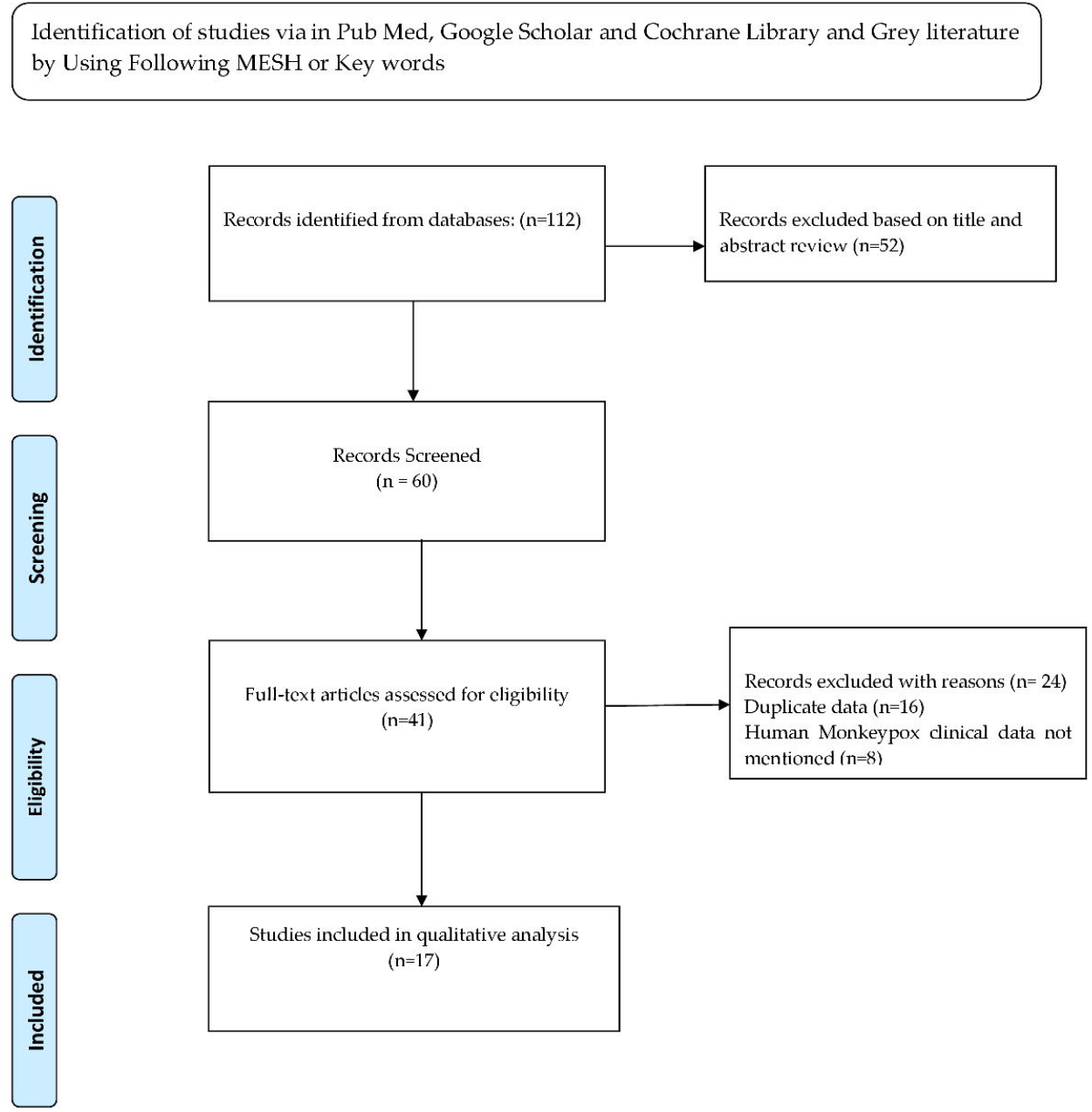
PRISMA Flow Diagram for the study selection.

### Epidemiology, Clinical Characteristics, Transmissibility and Diagnosis of Human MPX infection in the previous Outbreaks

The monkeypox virus first replicates at the site of inoculation, spreads to nearby lymph nodes, and causes viremia during the incubation period, which lasts for roughly 7 to 14 days. Thereafter, a prodromic stage is characterized by fever, chills, headache, sweats, fatigue, sore throat, muscle ache, and lymphadenopathy [4-5]. However, all patients may not report initial prodromal illness and it may start as a rash with lesions progressing through several stages [4-5]. Commonly, the prodromal stage may last for five days, followed by a skin eruption phase which usually manifests within one to four days of the appearance of fever and lymphadenopathy and is characterized by a few to several thousand lesions as 2 to 5 mm diameter macules and continues for a period of two to three weeks [4-5]. Secondary bacterial infection and dehydration are more common due to the disruption of skin from the lesions [4]. Lymphadenopathy occurring in the early stage of the illness is a typical sign of monkeypox differentiating human monkeypox from smallpox and chickenpox [5]. The rash associated with monkeypox is often described as painful, but in the healing phase or crusts it can become itchy and the rash develops from macules through papules, vesicles, pustules, and ultimately crusts [6]. Gastrointestinal symptoms are also often experienced by patients with MPX including, diarrhoea, vomiting, decreased appetite and dysphagia [7]. In addition, most commonly MPX infections affect the eyes causing secondary bacterial infections leading to swollen and red eyes, sensitivity to light and in severe cases it may lead to loss of vision [8]. In some cases, the respiratory tract is also affected causing severe cough, difficulty in breathing or bronchopneumonia [8]. In severe cases, serious complications such as encephalitis and sepsis are common causes of mortality [9]. Monkeypox case fatality rates vary depending on the virus lineage and range from 1% to 11%., but among the youngest children and immunocompromised patients, it was high (15%) [1-2, 5].

The human-to-human transmission was high with the Congo Basin clade and less with the West African clade, but the outbreaks of the West African clade in Nigeria have shown a high transmission between close contacts than initially reported [1-2]. In addition to direct contact with lesion material or body fluids, person-to-person transmission can also occur through large respiratory droplets after prolonged face-to-face contact [10-11]. Virus-contaminated materials, including mattresses and clothing, can also transmit the disease [12]. There have been reports of the MPXV spreading from the affected mother to the fetus [13]. Case studies of miscarriage and fetal death with the stillborn showing diffuse cutaneous maculopapular skin lesions exist [14].

Although serological and antigen detection tests can be useful when no virologic specimen is available, still they cannot offer a conclusive diagnosis [15]. Serologic testing identifies anti-orthopoxvirus IgG and/or IgM antibodies in the absence of recent vaccination [15]. The MPXV DNA identification by Reverse transcriptase polymerase chain reaction (RTPCR) on lesion specimens is the definitive diagnostic test for MPX infection [14]. The most specific specimen used for the diagnosis of the MPX includes two samples of lesion swabs based on the disease phase [15]. Monkeypox patients frequently have abnormal aminotransferases, leukocytosis, thrombocytopenia, and hypoalbuminemia in their laboratory tests. Acute inflammation, significant spongiosis, keratinocyte ballooning degeneration, and cutaneous oedema can all be associated with various viral infections, according to histopathological examination [15].

### Epidemiology, Clinical Characteristics, Transmissibility and Diagnosis of Human MPX infection in the Current Monkeypox Outbreak 2022

The published and grey literature related to the present outbreak yielded a total of 17 studies (n=17,811) from 1^st^ May 2022 to December 2022. Predominantly, most of the studies are from the Europe continent, where the monkeypox virus was not endemic. In the present outbreak of 2022, the transmission of MPXV is between person-to-person and has intricated epidemic in several nations to date. The median age group infected in the current outbreak was 30-40 years young adults and the majority were males [16-17, 20-29, 40, 56,59,61-64]. So far, in the MPX outbreak 2022, most of the reported cases but not exclusively have been identified in men who have recent sexual contact with new or multiple male partners [1]. The primary risk factor for MPXV transmission is thought to be sexual contact [13,18-19]. In some cases of young children, household transmission has also occurred. Typical symptoms seen in the present MPX outbreak include rashes at various sites of the body anogenital lesions, mucosal lesions, fever, lethargy, myalgia, headache, lymphadenopathy (cervical, axillary, Inguinal), influenza-like illness, sore throat, nasal congestion, cough, proctitis, tonsillitis, bacterial skin abscess, vesicular-umbilical and pseudo-pustular skin lesions, asthenia, exanthema, skin lesions in the arm, perianal and trunk area, genital area, chest and legs, wrist, pectoral, fingers, hand, ulcerated ventral tongue, genital vesicles, odynophagia, perianal Abscess, tonsillar abscess, sore throat, rectal pain with rectal perforation, Penile oedema[20-29,40,56,59,61-64]. There are many atypical clinical symptoms observed in the current Monkeypox Outbreak 2022 such as, not all patients reported initial prodromal febrile illness and it started as a rash appearing as pimples or blisters and painful or itchy with lesions progressing through several stages including scabs before healing or only flu-like symptoms or only lesions in the anogenital region and the mouth, hands, palms and alimentary tract areas, rectal pain and penile oedema without any prodromal symptoms[18-29,40,56]. In a case series of 2022 outbreaks, study findings have shown that the MPXV DNA was found in semen samples, which was unknown in previous reports that the virus can spread through vaginal or semen fluids [18, 29]. Patients are contagious as soon as their illness develops., and the infectious lesions comprise viruses throughout disease phases until all skin lesions are scraped over and reepithelialization has occurred, which can take up to 4 weeks [17]. The mean incubation period was 7 (3-21) days [20-29]. For the confirmation of monkeypox virus diagnosis, Swab specimens of the skin or genital lesions, exudate and/or crusts from lesions in the palms, throat or nasopharyngeal, semen, urine and faeces and blood by using Reverse transcriptase polymerase chain reaction (RTPCR) test [20-29, 40, 56,59,61-64]. Studies have also shown that monkeypox DNA was detected in the saliva, vaginal and seminal fluid of infected patients [20]. Surprisingly in the present outbreak, DNA of MPXV was also found in the anal or oropharyngeal swabs of asymptomatic MSM [60-61].

Patients hospitalized for the treatment of severe painful bacterial anogenital infections [20-29, 40,56,59,61-64]. The majority were suffering from human immunodeficiency virus (HIV) and other sexually transmitted diseases (STDs) [20-29, 40, 56,59,61-64]. The majority of patients were hospitalized due to concomitant severe bacterial infections and rectal pain, and proctitis and were treated for secondary bacterial infections with antimicrobials, and supportive treatment including anti-inflammatory, and antihistaminic medications. The most common antiviral agents used for the treatment of MPX infection include oral tecovirmat, brincidofovir and topical cidofovir (eye involvement). In the present outbreak, the case fatality rate was 3-6% [1]. However, there were two deaths reported in the presentation including studies. Overall, when compared to previously documented outbreak reports in endemic west and central African regions in the past 30 years, the clinical pattern of the monkeypox cases in non-endemic regions of the present outbreak has been completely atypical, varying in MSM and the manifestations among infected. Unexpectedly, cases with no symptoms had MPXV DNA positive tests as well. In contrast to outbreaks in endemic locations, there is a higher rate of hospitalization for the management of pain and necrotizing secondary bacterial infection in the anorectal region. The majority of the reported MPXV cases were associated with sexual contact only and the viral DNA has been identified even in seminal, anogenital and saliva samples along with swabs from skin lesions. **Table 1 Presents Clinical Studies of the Monkeypox Outbreak 2022** [20-29, 40, 56,59,61-64].

**Table 1.**
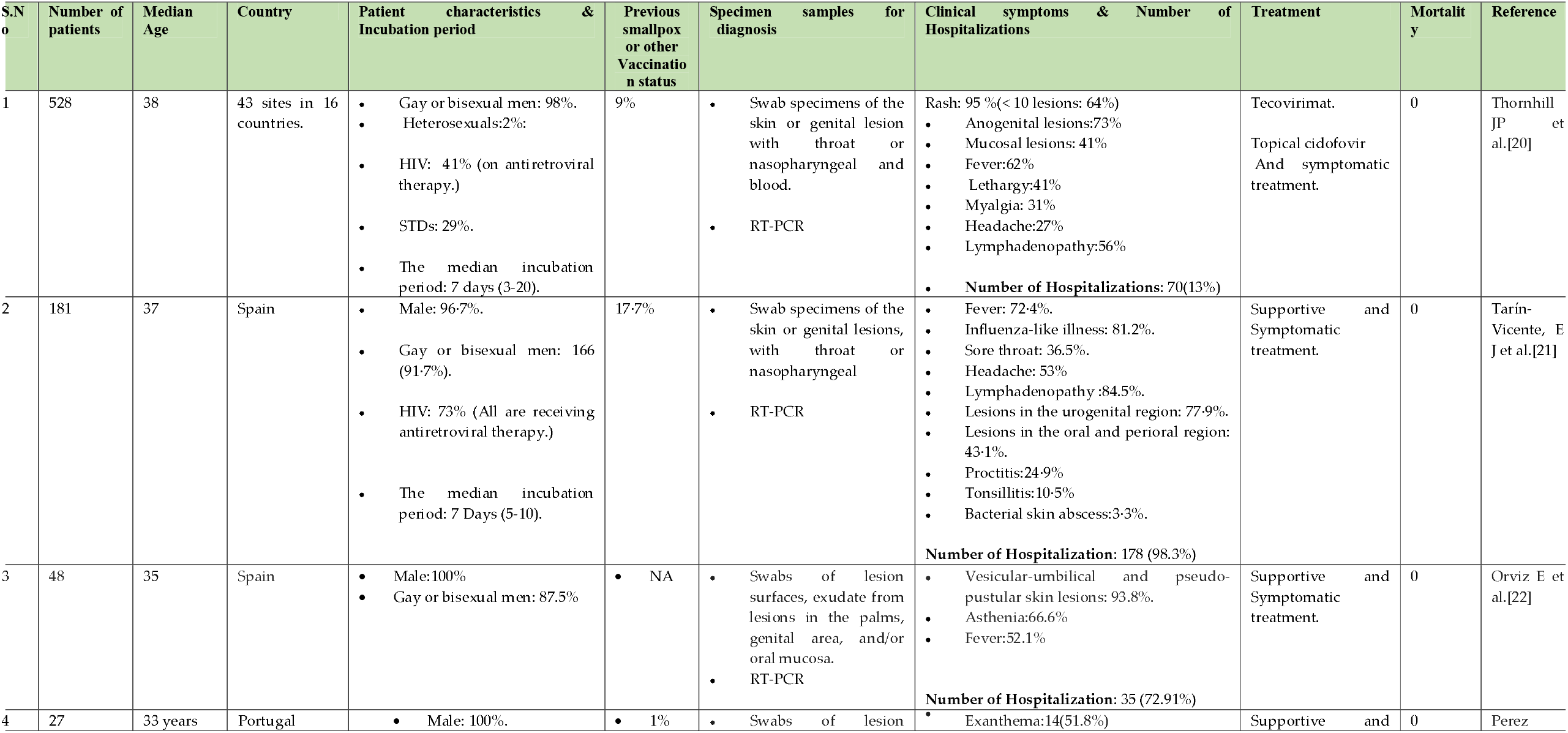
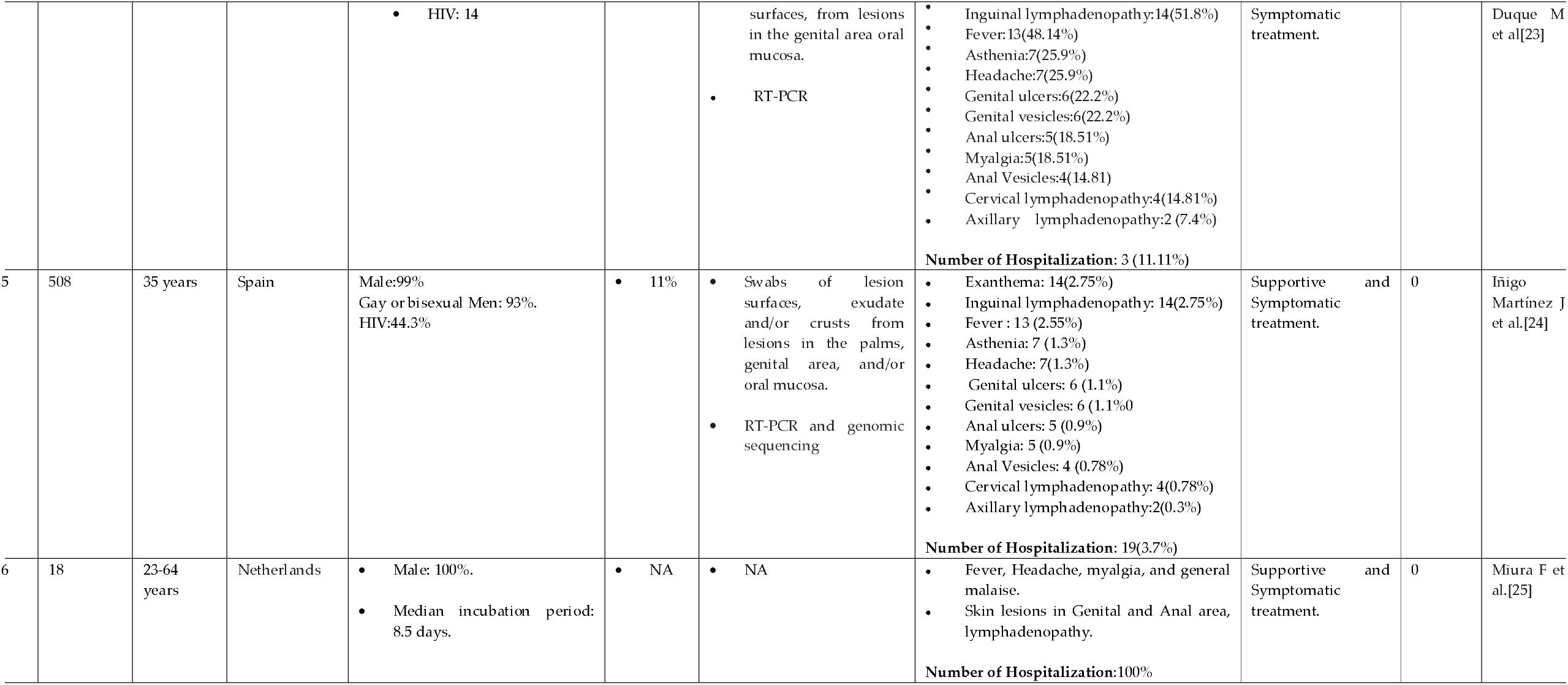
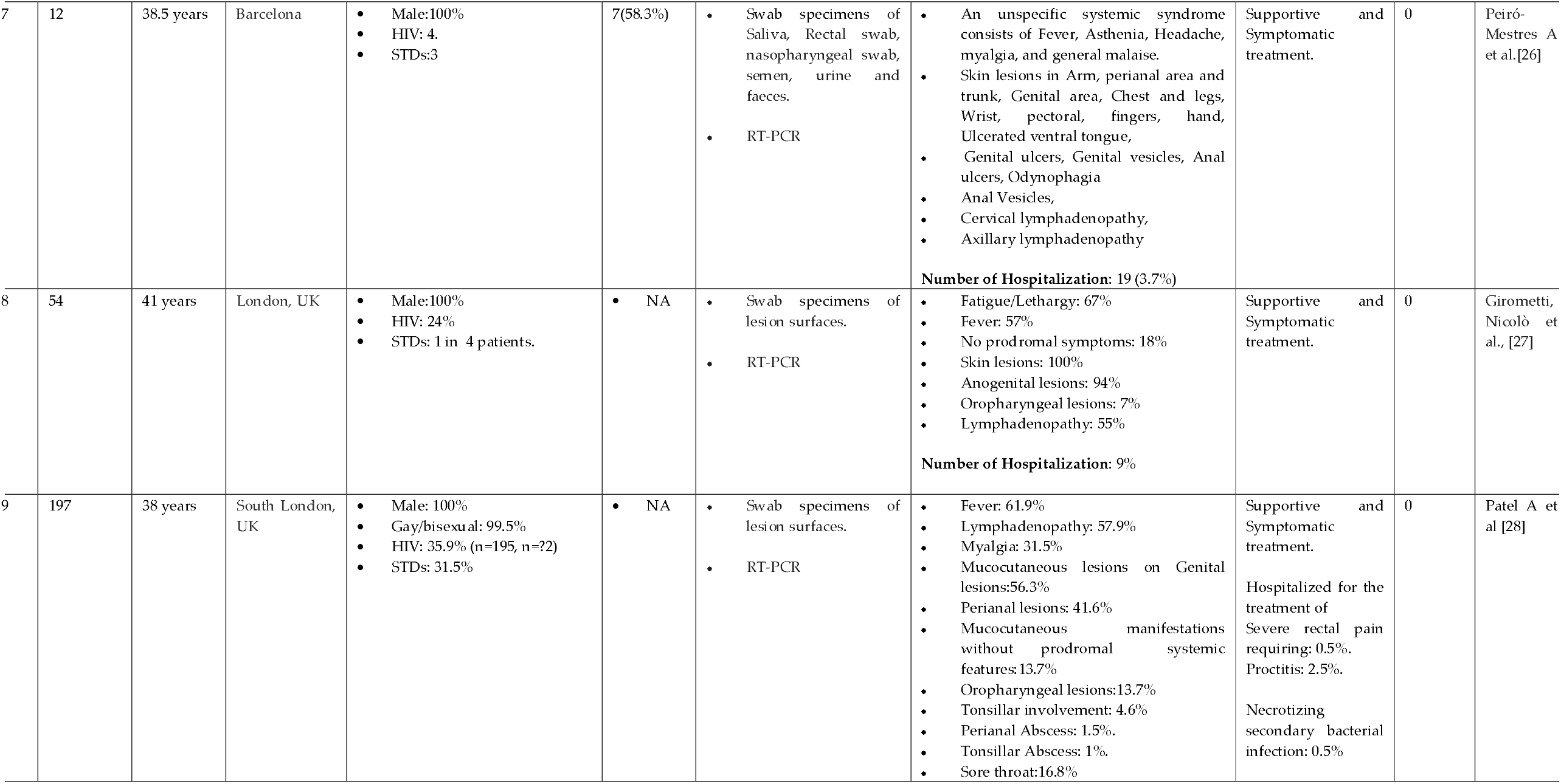
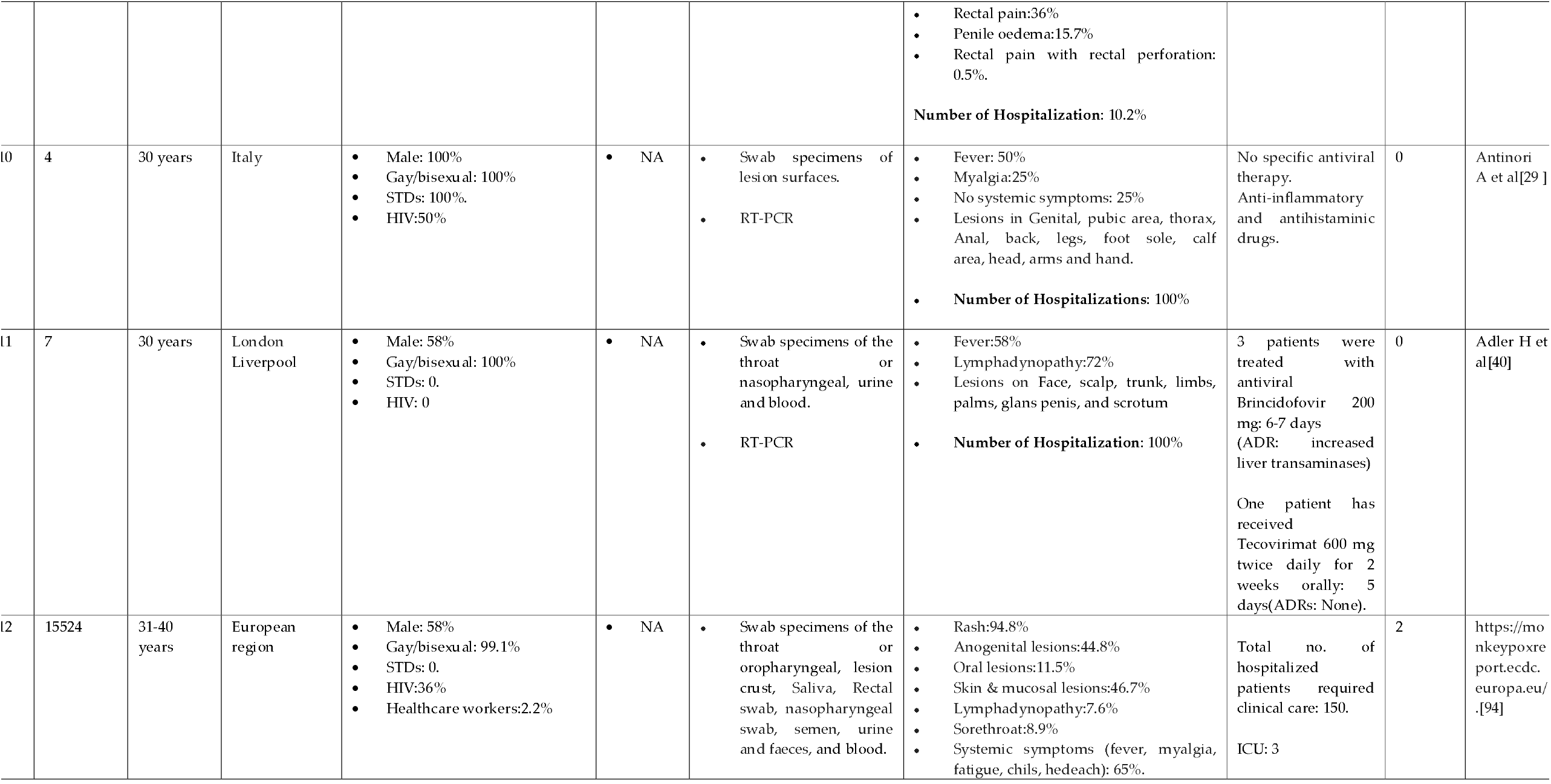
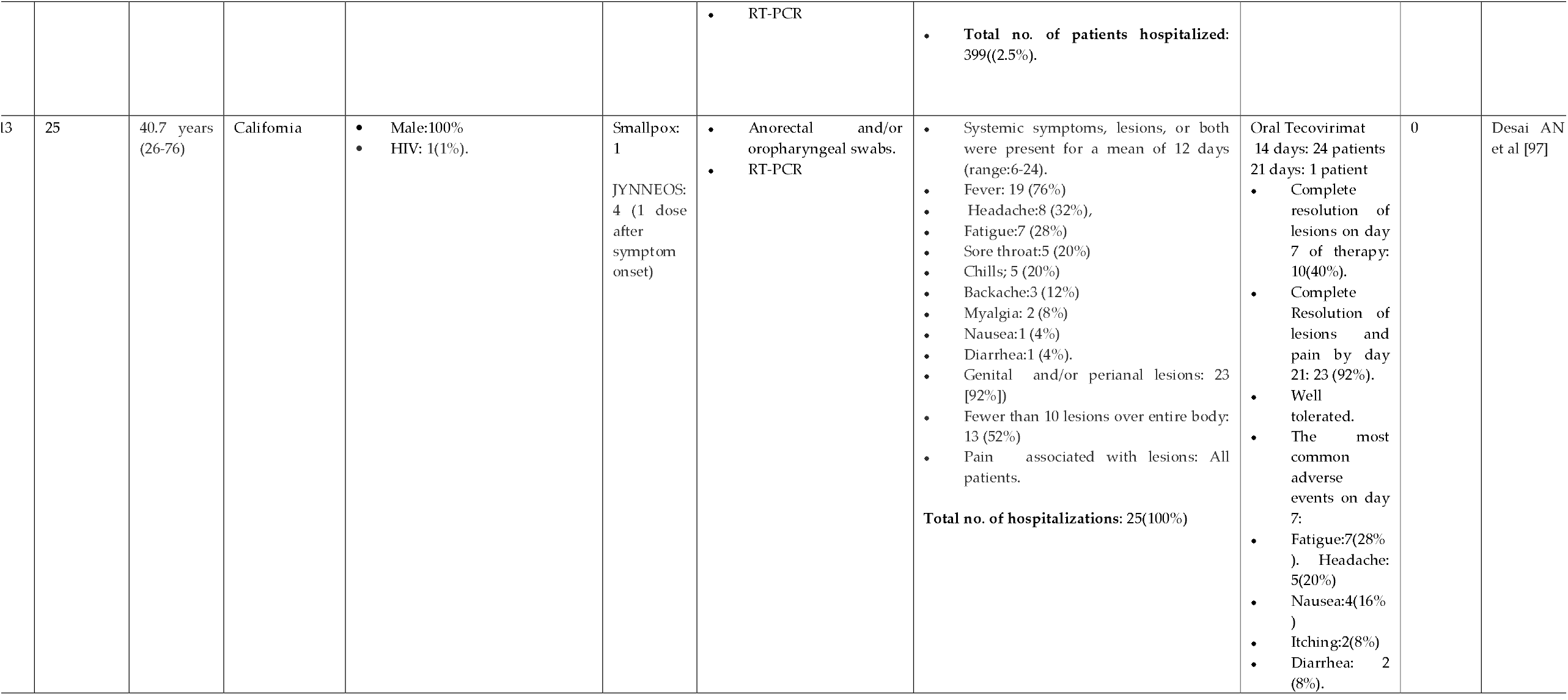
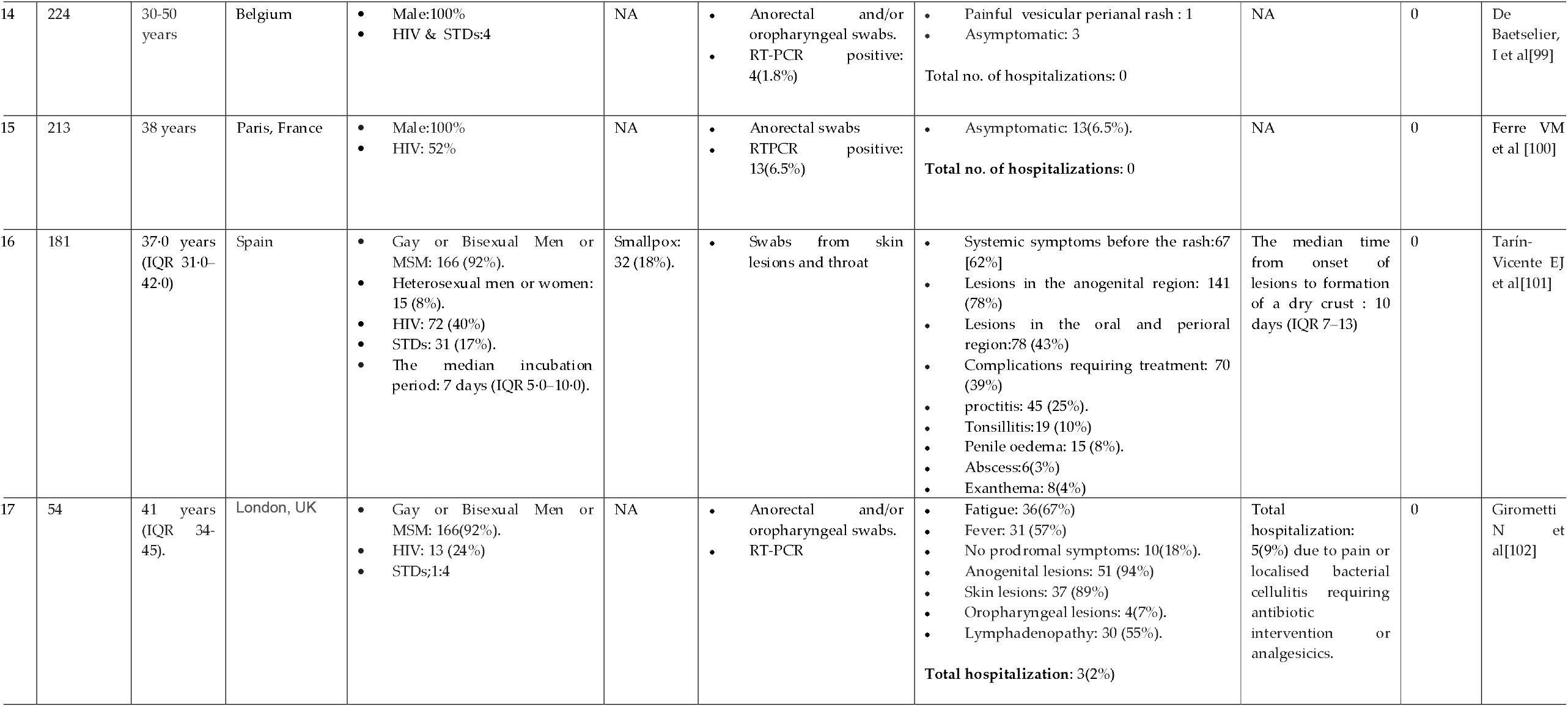
Clinical studies of 2022 Monkeypox outbreak. MPX:Monkeypox ; HIV: Human Immuno Deficiency ; STD: Sexually Transmitted disease: RTPCR: reverse transcriptase polymerase chain reaction. MSM: Men who have sex with men ; NA: Not Available.

### Current Preventive and Treatment Strategies

Based on the findings of earlier studies from countries where monkeypox is endemic, patients with MPX may benefit from medical supportive treatment to lessen the effects of lesions, which comprises preventing and treating secondary complications, maintaining sufficient hydration and nutrition, and safeguarding susceptible anatomical areas like the eyes and genitalia [30-31].Currently, only one antiviral agent, tecovirmat is approved by the United States Food and Drug Administration (USFDA) for the treatment of human smallpox disease caused by the *Variola virus* in adults and children [32-33]. However, tecovirimat is being made available through the United States Centre for Disease Control (USCDC) under an FDA authority called Expanded Access or “compassionate use.” There are currently no human data demonstrating the efficacy of tecovirimat for the treatment of monkeypox, or the safety and pharmacokinetic profile. While there are no MPXV-specific antivirals currently approved, the US CDC and WHO Interim Clinical Guidelines recommended three antiviral agents such as tecovirimat, cidofovir and brincidofovir for the treatment of MPX infection, have been made available to treat monkeypox via an expanded access Investigational New Drug (IND) protocol [32-33]. According to the CDC and WHO interim guidelines, the aforementioned antiviral agents should be considered only in patients with monkeypox with severe disease and those at risk for severe diseases such as pregnant or breastfeeding women, patients younger than eight years of age, patients with complications of the infection and in patients with monkeypox caused by accidental implantation in the eyes, mouth, or other areas along with supportive treatment [32-33]. There are oral capsules and injectable forms of Tecovirimat that can be used intravenously [32-33]. To increase oral absorption, tecovirimat should be given along with a high-fat meal [32-33]. Currently, there are several clinical trials of tecovirmat are underway or planned (PALM-007, PLATINUM, WHO/ARNS, and ACTG5418) all over the world to provide required data on the safety and efficacy of monkeypox infection [32-33]. A recent case series of 25 patients with confirmed MPX infections who received a course of tecovirmat therapy for 14 days (one patient for 21 days) on weight-based, for every 8 or 12 hours, on compassionate use basis concluded that oral treatment with tecovirimat was well tolerated, with minimal adverse drug reactions [59].

Vaccinia Immune Globulin Intravenous (VIGIV), a hyperimmune globulin is also indicated by the US CDC guidelines, particularly in individuals with immunodeficiencies or individuals for whom vaccination is contraindicated [32-34]. If monkeypox lesions involve the eyes (conjunctivitis and oedema of the eyelids, focal lesions on the conjunctiva and along the margins of the eyelids) off-label use of trifluridine or vidarabine eye drops or ointments applied every four hours for 7 to 10 days with or without intravenous VIG are indicated [31,34-36]. However, topical corticosteroids to control inflammation may lead to virus persistence and prolonged corneal damage, hence steroids should not be used for treating monkeypox involving the eyes [33-34]. Enhanced lubrication or topical antibiotics could be considered for monkeypox involving the eyes [31, 35]. To avoid long-term ocular problems such as chronic pain, corneal opacity, scar tissue, and permanent loss of vision, an early therapy to enhance virus elimination may be taken into consideration in MPX patients [36-55]-

Evidence from the literature has shown that the smallpox vaccine protects 85% against monkeypox via cross-immunity [36]. There are three available vaccines for orthopoxvirus: ACAM2000, MVA-BN7 or JYNNEOS (Imvamune or Imvanex) and LC16 [32, 37-38, 50]. However, no data is available yet on the effectiveness of these vaccines in the current outbreak. The first-generation vaccines are smallpox vaccines produced by propagated and harvested primarily from the skin of live animals, infecting the abdomen and flanks of calves or other large animals (e.g., calf, sheep, water buffalo lymph) or chorioallantois membrane (CAM) of embryonated hens’ eggs during the intensified smallpox eradication program consist of live vaccinia virus [36, 55]. However, the same *Vaccinia virus* strains used to produce the first-generation vaccinations are used to produce the second-generation smallpox vaccines (ACAM2000) using designated cell lines, or clonal virus strains plaque-purified from conventional vaccine stocks [50,55]. Later, the third-generation vaccines were produced and specifically developed as safer vaccines towards the end of the eradication phase using the more attenuated smallpox vaccine strains by a further passage in cell culture or animals (LC16 and MVA-BN) [55]. In the year 2019, MVA-BN (JYNNEOS) was approved for the prevention of smallpox and monkeypox by the US FDA followed by national regulatory authorities such as US CDC and the European Medicines Agency (EMA) [52, 54-55]. However, due to the limited supply of vaccine JYNNEOS, the USFDA issued an Emergency Use Authorization (EUA) on August 9, 2022, to administer the vaccine by intradermal injection for individuals ≥ 18 years to those at high risk for monkeypox infection [60].

In collaboration with local public health authorities, the Advisory Committee on Immunization Practices (ACIP) recommends the use of either the ACAM 2000 or MVA vaccine for specific health professionals at increased risk for occupational exposure to MPXV, including laboratory personnel conducting screening tests for Orth poxviruses [32]. For certain populations, such as toddlers, women who are pregnant or nursing, or those who have immune suppression, the monkeypox vaccine is typically not advised as a pre-exposure prophylaxis [32, 55]. After carefully weighing the risks and benefits, vaccination for monkeypox as post-exposure prophylaxis may be recommended for some demographic groups, such as those who are pregnant, have young children or have immune suppression, such as those who have HIV [32,55]. Individuals with a greater or intermediate risk of MPX contact and/or eligible for post-exposure prophylaxis should be vaccinated within four days of exposure [32,55]. According to the CDC guidelines, a high-risk exposure is considered as either “unprotected contact between a person’s skin or mucous membranes and the skin, lesions, or bodily fluids from a person with monkeypox (sexual contact, inadvertent splashes of patient saliva to the eyes or oral cavity of a person, ungloved contact with the patient) or contaminated materials(clothing)” or “being inside the patient’s room or within six feet of a patient during any procedures that may create aerosols from oral secretions, skin lesions, or resuspension of dried exudates without wearing an N95 or equivalent respirator and eye protection”. Intermediate risk exposure considered as either “being within six feet for three hours or more of an unmasked person with monkeypox without wearing, at a minimum, a surgical mask” or “engaging in an activity that results in contact between sleeves and other parts of an individual’s clothing and the patient’s skin lesions or bodily fluids or their soiled linens or dressings (turning, bathing, or assisting with transfer) while wearing gloves but not wearing a gown” [32]. People who have been exposed to the MPXV and haven’t had the smallpox vaccine in the past three years should think about receiving the vaccine [32]. ACAM2000 may be administered via a procedure called scarification for the indicated. Although vaccination can be considered for exposures lasting up to 14 days if administered between days 4 and 14, it is believed to lessen disease symptoms but not to prevent them. ACAM2000 is administered 4 weeks following the second dose of MVA-BN, at which point a person is deemed fully immunized[32]. Nevertheless, even after receiving the vaccine, individuals should keep taking precautions to prevent disease by preventing close, skin-to-skin contact, particularly intimate contact, with a person who has monkeypox [32,55].

## Discussion & Conclusions

The most recent pandemic, COVID-19, reminds the need to constantly be prepared for new virus outbreaks. Viruses that haven’t been observed in a while resurface. Currently, the monkeypox outbreak is a new viral infection threatening all healthcare authorities in the world because of the increasing number of cases in nonendemic regions of the world. However, despite the possibility of a new pandemic at any moment, new antiviral drugs and vaccines should be there to control the virus’s severity and mortality. Because of variability in clinical features of the monkeypox outbreak 2022, such as lesions in the anogenital region and the mouth, hands and palms before (or without) other prodrome symptoms, rectal pain and penile oedema, myalgia, lethargy, sore throat and asymptomatic cases as well. These novel clinical presentations should be included in public health messaging to aid early diagnosis and reduce onward transmission. Hence, due to the increased rate of hospitalization including intensive care unit admission and slowly rising fatality rate, clinicians must be vigilant to test and trace those patients who present with these atypical symptoms and asymptomatic high-risk category population, predominantly heterosexuals and or men who have sex with men.

## Data Availability

All data produced in the present study are available upon reasonable request to the authors

## Acknowledgement

None.

## Data Availability

The data used in this systematic review is available from the corresponding author with a reasonable request.

## Author Contributions

All authors contributed to the literature search, data extraction and draft of the manuscript. All authors reviewed and approved the final version of the manuscript.

